# Identification of clinically-relevant genetic alterations in uveal melanoma using RNA sequencing

**DOI:** 10.1101/2023.12.03.23299340

**Authors:** R.J. Nell, M. Versluis, D. Cats, H. Mei, R.M. Verdijk, W.G.M. Kroes, G.P.M. Luyten, M.J. Jager, P.A. van der Velden

**Affiliations:** Department of Ophthalmology, Leiden University Medical Center, Leiden, the Netherlands; Department of Pathology, Leiden University Medical Center, Leiden, the Netherlands; Department of Pathology, Erasmus University Medical Center, Rotterdam, the Netherlands; Department of Biomedical Data Sciences, Leiden University Medical Center, Leiden, the Netherlands; Department of Clinical Genetics, Leiden University Medical Center, Leiden, the Netherlands

**Author notes:** Corresponding author, Department of Ophthalmology, Leiden University Medical Center, PO Box 9600, 2300 RC Leiden, The Netherlands.

## Abstract

**Introduction:** Uveal melanoma is a lethal intraocular tumour, in which the presence of certain genetic alterations correlates with the risk of metastatic dissemination and patient survival. RNA data is typically used to transcriptionally characterise tumours and their micro-environment. In this study, we tested the detectability of all key genetic alterations in uveal melanoma from RNA sequencing data.

**Methods:** Cohort-wide gene expression profiling was used to classify tumours at the transcriptional level. In individual samples, copy number alterations affecting chromosomes 3 and 8q were analysed by measuring expressed allelic imbalances of heterozygous common single nucleotide polymorphisms. Mutations in *GNAQ, GNA11, CYSLTR2, PLCB4, BAP1, SF3B1* and *EIF1AX* were identified by screening of hotspot regions and by evaluating their transcriptional effects. All findings were cross-validated with DNA-derived data in a training cohort of 80 primary uveal melanomas studied by The Cancer Genome Atlas (TCGA) initiative, and in five prospectively analysed cases from our institution.

**Results:** Unsupervised gene expression profiling strongly correlated to the presence of chromosome 3 alterations, but was not reliable in identifying other (clinically-)relevant genetic alterations. However, the presence of both chromosome 3 and 8q copy number alterations could be successfully inferred from expressed allelic imbalances in most tumours. The majority of mutations were adequately recognised at the RNA level by their nucleotide changes (all genes), alternative splicing around the mutant position (*BAP1*) and transcriptome-wide aberrant splice junction usage (*SF3B1*). Notably, in the TCGA cohort we detected previously unreported mutations in *BAP1* (n=3) and *EIF1AX* (n=5), that were missed by the original DNA sequencing. In our prospective cohort, all mutations and copy number alterations were successfully identified at the RNA level by combining the described approaches.

**Conclusion:** In addition to providing gene expression levels and profiles, RNA from uveal melanomas presents insights into the expressed tumour genotype and its phenotypic consequences. Such complete analysis of transcriptional data may augment or even substitute current DNA-based approaches, and has potential applicability in both research and clinical practice.

## Introduction

Uveal melanoma is a lethal intraocular tumour characterised by a high metastatic rate and limited treatment options once disseminated [1]. In contrast to most other malignancies, it presents as a relatively simple genetic disease with a low mutational burden and limited number of structural variants [2, 3]. Practically all tumours harbour a mutually-exclusive mutation in *GNAQ, GNA11, CYSLTR2* or *PLCB4*, which activates the Gα_q_ signalling pathway [4-7]. Most uveal melanomas also carry a so-called BSE (*BAP1, SF3B1* or *EIF1AX)* mutation [2, 8-10]. Copy number losses of chromosome 3 and increases of chromosome 8q are the most prevalent structural alterations, and occur in specific combinations together with the BSE mutations. These various molecular subtypes have been associated with distinct gene expression profiles (GEP) and are differentially correlated to the risk of early metastatic dissemination and consequent patient survival (**Table 1**).

**Table 1.** Summary of three most typical combinations of GEP classification, mutations, chromosomal copy number alterations and their correlation to the risk of (early) metastatic dissemination [1].

| GEP | Mutations | Copy number alterations | Risk to (early) metastasis |
| --- | --- | --- | --- |
| Class I | Gα<sub>q</sub> + *EIF1AX* |  | Low |
|  | Gα<sub>q</sub> + *SF3B1* | +8q | Intermediate |
| Class II | Gα<sub>q</sub> + *BAP1* | −3, +8q | High |

The prognostic value of genetic alterations in uveal melanoma has been recognised for more than two decades. Traditionally, copy number alterations were studied using cytogenetic techniques, such as karyotyping, fluorescence in situ hybridisation, multiplex ligation-dependent probe amplification and array genotyping [11-15]. Technical advancements enabled genome-wide analyses by next-generation sequencing and led to the discovery of recurrent mutations [2, 8-10]. Nowadays, (targeted) sequencing can be used to detect all relevant alterations – both copy number alterations and mutations – in one single analysis [16, 17].

In addition to these DNA-based approaches, several studies have identified clinically-relevant transcriptional subtypes of uveal melanoma [18-21]. Most commonly, tumours are divided into class I or II based on their bulk GEP. This classification can be established using dimensionality reduction and sample clustering techniques of total RNA expression data, or by evaluating expression variation in a selection of genes. Intriguingly, this transcriptional classification of tumours is known to strongly correlate with the presence of genetic alterations affecting chromosome 3 (**Table 1**). Given that part of the bulk GEP may be derived from admixed non-malignant cells, such as tumour-infiltrating immune cells, RNA data can also provide insights into the tumour microenvironment of uveal melanomas [22, 23].

More recently, the presence and consequences of genetic alterations at the transcriptional level have gained interest in the context of uveal melanoma. Mutations in *SF3B1*, a gene encoding for a core component of the cell’s splicing machinery, were found to lead to a recurrent profile of aberrant splice junction usage [2, 24]. As another example, in three large-scale genomic studies, a variety of complex *BAP1* alterations were only identified after using RNA sequencing data in addition to DNA-derived data [21, 25, 26]. Finally, *CYSLTR2* mutant uveal melanomas recurrently showed silencing of the wild-type allele and preferential expression of the p.L129Q mutation [27]. These examples emphasise the value of investigating RNA in addition to DNA to identify the presence and consequences of certain genetic alterations.

In this study, we apply various bioinformatic approaches to detect all key genetic alterations in uveal melanoma by only using tumour-derived RNA sequencing data. Cohort-wide GEP is used to identify class I and II tumours. In individual samples, copy number alterations are analysed by measuring expressed allelic imbalances of heterozygous common single nucleotide polymorphisms (SNPs). Mutations are identified by screening of hotspot regions and by evaluating their transcriptional effects. Our findings are cross-validated with DNA-derived data in a training cohort of 80 primary uveal melanomas originally studied by The Cancer Genome Atlas (TCGA) initiative, and in five prospectively analysed cases from our institution.

## Methods

### Collection and analysis of retrospective cohort

RNA sequencing files from the 80 primary uveal melanomas studied by the TCGA [13] were downloaded from the NCI Genomic Data Commons data portal (GDC; https://portal.gdc.cancer.gov). After reconversion to FASTQ files using samtools (version 1.1), the reads were aligned using STAR (version 2.5.3a) to human reference genome GRCh38. Gene counts were generated using htseq-count (version 0.6.0) and annotated with Ensembl (version 87). DESeq2 (version 1.30.0) was used for variance stabilising transformation of the count data, which formed the input for the uniform manifold approximation and projection (UMAP) analysis. An expression signature of 200 genes characterising the two visually separated clusters was identified using ClaNC, and was used to classify tumours as ‘class I’ or ‘class II’ based on the relative expression of chromosome 3 genes (i.e. high or low, respectively).

The aligned RNA sequencing files were analysed for variants using VarScan (version 2.3), which were annotated using snpEff/snpSift (version 5.1) according to dbSNP (version 151). Only heterozygously and highly expressed common SNPs (flagged as ‘common’, total number of reads > 40, number of reference reads > 2, number of alternate reads > 2) were included in the downstream analysis. For all SNPs, the fractions alternate reads of total reads were visualised according to their chromosomal positions. Segmentation was performed manually. RNA-derived allelic (im)balances were compared to DNA-measured copy number information determined from Affymetrix SNP 6.0 arrays, available from the Pan-Cancer Atlas initiative [28].

All aligned RNA sequencing files were further screened for (hotspot) mutations in *GNAQ* (p.Q209, p.R183 and p.G48), *GNA11* (p.Q209, p.R183), *CYSLTR2* (p.L129), *PLCB4* (p.D630), *EIF1AX* (entire gene), *SF3B1* (exon 14) and *BAP1* (entire gene) using freebayes (version 1.3.6), and via manual inspection using the Integrative Genomics Viewer (version 2.8.10). The presence of mutations was compared to those detectable in aligned DNA sequencing files generated by exome-captured sequencing (n=80) or low-pass whole-genome sequencing (n=51) downloaded from the GDC data portal.

The bioinformatic analyses were carried out using R (version 4.0.3) and RStudio (version 1.4.1103). All custom scripts are available via https://github.com/rjnell/um-rna.

### Collection, isolation and analysis of prospective cohort

Five snap-frozen primary uveal melanoma specimens were collected from the biobank of the Department of Ophthalmology, Leiden University Medical Center (LUMC). These samples were originally obtained from patients treated by an enucleation in the LUMC. This study was approved by the LUMC Biobank Committee and Medisch Ethische Toetsingscommissie under numbers B14.003/SH/sh and B20.026/KB/kb.

RNA from all specimens (25x 20μm sections) was isolated using the QIAamp RNeasy Mini Kit (Qiagen, Hilden, Germany), following the manufacturer’s instructions. 100 ng of RNA per sample was sequenced by GenomeScan (Leiden, the Netherlands). Sample preparation was performed using the NEBNext Ultra Directional RNA Library Prep Kit for Illumina (New England Biolabs, Ipswich, USA) according to the NEB #E7240S/L protocol. In short, mRNA was isolated from total RNA using oligo-dT magnetic beads. After fragmentation of the mRNA, cDNA synthesis was performed and used for ligation with the sequencing adapters and PCR amplification. Quality and yield were measured with the Fragment Analyzer (Agilent, Santa Clara, USA). Next, clustering and DNA sequencing was performed using the cBot and HiSeq 4000 (Illumina, San Diego, USA) according to the manufacturer’s protocols. HiSeq control and image analysis, base calling and quality check were carried out using HCS (version 3.4.0), the Illumina data analysis pipeline RTA (version 2.7.7) and bcl2fastq (version 2.17). The raw sequencing files underwent the same alignment and analysis pipeline as the TCGA data files to perform GEP clustering and infer copy number alterations and mutations.

DNA from uveal melanomas (25x 20μm sections) was isolated with the QIAmp DNA Mini Kit (Qiagen) according to the manufacturer’s instructions. Digital PCR was performed to measure the copy number values of chromosome 3 and 8q and to confirm mutations in *GNAQ, GNA11, EIF1AX* and *SF3B1*. These experiments were carried out using the QX200 Droplet Digital PCR System (Bio-Rad Laboratories, Hercules, USA) following established protocols [22, 27, 29-31], and two new assays listed in **Supplementary Table 1**. The presence and pathogenicity of *BAP1* alterations was confirmed by targeted Sanger or next-generation sequencing and an immunohistochemical staining, both performed as routine diagnostic analyses in two ISO accredited laboratories (Departments of Clinical Genetics and Pathology, LUMC) [16, 32].

## Results

### Estimation of genetic alterations via cohort-wide gene expression profiling

The RNA-derived classification of primary uveal melanomas into GEP class I or II is originally based on unsupervised clustering of transcriptome-wide expression data into two groups [19, 20]. Concordantly, a dimensionality reduction analysis (UMAP) successfully divided the TCGA tumours (n=80) into two main clusters based on their total transcriptional diversity (**Figure 1A**). To investigate expression patterns underlying this subdivision, we inferred a signature of 200 genes characterising the two clusters. The largest proportion (82/200, 41%) of these genes were located on chromosome 3 (**Figure 1B and Supplementary Table 2**), with systematically lower levels of expression in one cluster of tumours (n=40, marked as GEP class II) relative to the other (n=40, marked as GEP class I). This classification almost completely overlapped with the true DNA-determined copy number status of chromosome 3: 40/40 GEP class II tumours demonstrated loss of this chromosome (i.e. monosomy, or isodisomy in polyploid melanomas), compared to 3/40 GEP class I tumours (**Figure 1C and Supplementary Table 3**). The three discordant cases were the only tumours showing arm-level variation of chromosome 3 loss (n=2) or an unusual trisomy 3 in the context of polyploidy (n=1, **Supplementary Figure 1A and B**). In line with the frequent co-occurrence of chromosome 3 copy number losses and *BAP1* alterations, mutations in *BAP1* were present in 40/40 GEP class II and 0/40 GEP class I tumours (**Figure 1C and Supplementary Table 3**).

**Figure 1.**
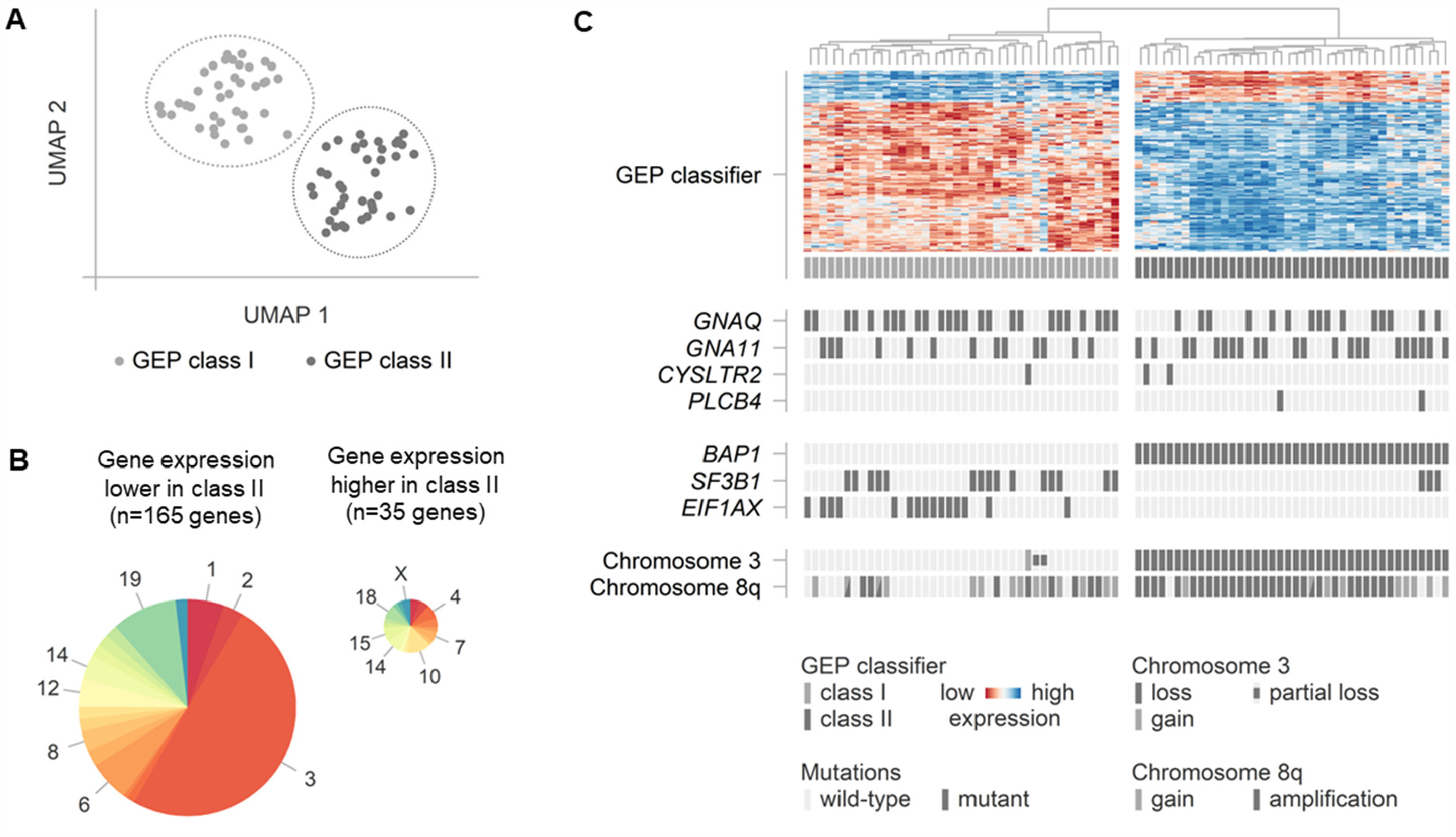
Cohort-wide gene expression profiling (GEP) to estimate the presence of genetic alterations in our training cohort of uveal melanomas studied by The Cancer Genome Atlas (n=80). (**A**) Two-dimensional uniform manifold approximation and projection (UMAP) analysis to identify two clusters of tumours: GEP class I and II. (**B**) Chromosomal location of signature genes characterising the two clusters. (**C**) Relative gene expression levels of signature genes characterising the two clusters in relation to the genetic alterations present per individual tumour.

In contrast, cohort-wide GEP clustering did not specifically overlap with the presence of chromosome 8q copy number changes or other mutations, even not after specifying the analysis to class I or II tumours only (**Figure 1C and Supplementary Figure 2**). These observations suggest that alternative approaches are needed to infer all (clinically-)relevant genetic alterations from RNA data.

### Identification of copy number alterations via RNA-expressed allelic imbalances

To detect chromosomal copy number alterations in individual tumours, we proceeded by measuring regional allelic imbalances in the RNA sequencing data. For this goal, highly expressed heterozygous common SNPs were identified and their so-called B-allele fractions (BAFs) were visualised according to their chromosomal positions. These values refer to the measured expression of one allele compared to the total expression of both alleles. Under copy number neutral conditions, two alleles – and thus both SNP variants – are equally abundant, resulting in BAFs of ∼50% (**Figure 2A**). In contrast, a chromosomal loss, such as monosomy 3, typically leads to BAFs close to ∼0% and ∼100%, as it is caused by an (almost) complete loss of one of the two alleles (‘loss of heterozygosity’, **Figure 2B**). Using this approach, we were able to correctly identify disomic tumours (n=40, with preserved balanced expression) and tumours with (partial) loss of chromosome 3 (n=39, showing imbalanced expression) from the RNA sequencing data in 79/80 (99%) cases of the TCGA cohort (**Figure 2C and Supplementary Table 3**). The only conflicting case was the one presenting with an unusual trisomy 3 in the context of polyploidy, which was also characterised by an allelic imbalance (**Supplementary Figure 1B**).

**Figure 2.**
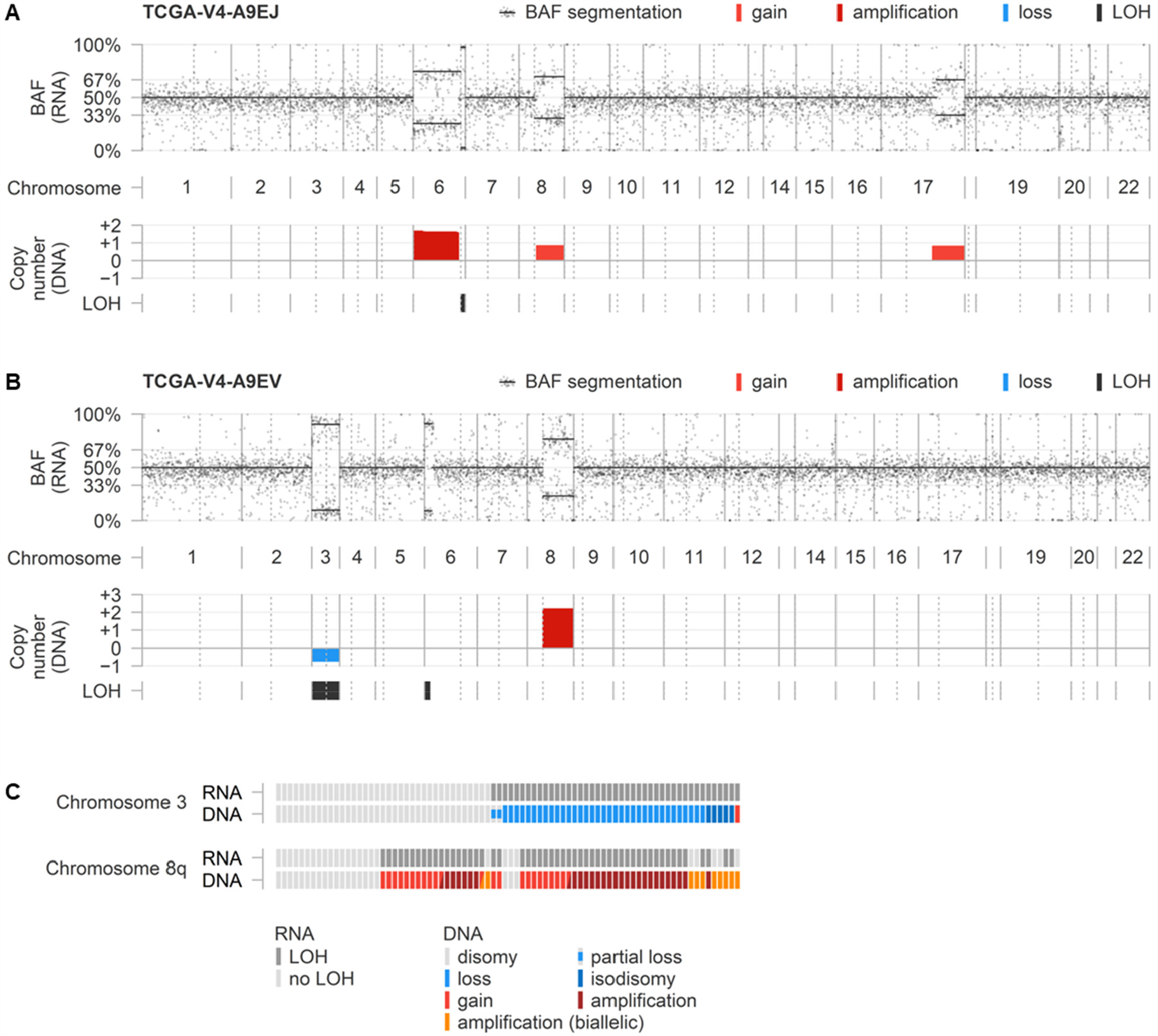
RNA-inferred allelic imbalances in relation to DNA-confirmed copy number alterations. (**A**) Example of a tumour showing balanced expression of chromosome 3, but imbalanced expression of chromosome 8q, in line with a disomy 3 and copy number alteration of chromosome 8q. Additionally, alterations affecting chromosome 6p, 6q and 17q (partial) are correctly identified. (**B**) Example of a tumour showing imbalanced expression of both chromosome 3 and 8q, in line with copy number alterations of the two chromosomes. Additionally, an alteration affecting chromosome 6p (partial) is correctly identified. (**C**) Overview of RNA- and DNA-derived observations in the 80 TCGA tumours.

This latter case illustrates that, similar to chromosomal losses, copy number increases typically also lead to allelic imbalances. A gain, such as of chromosome 8q (or chromosome 3 in the discordant case described above), is usually the result of an extra copy of one allele, leading to BAFs around ∼67% and ∼33% (**Figure 2A**). Larger amplifications mostly originate from several copies of one allele (with BAFs >67% and <33%, **Figure 2B**). On the other hand, amplifications derived from extra copies of both alleles or subclonal alterations result in BAFs closer to 50% and may be more difficult to recognise (**Supplementary Figure 1C**). In the TCGA cohort, chromosome 8q allelic imbalances were detected in the RNA sequencing data of 53 tumours, which all had a DNA-confirmed gain or amplification of chromosome 8q. The remaining 27 cases lacked measurable BAF deviations, and could be explained by a disomy 8q (n=21) or a copy number increase of both alleles (n=6). In aggregate, the presence or absence of any 8q alteration was correctly identified from RNA sequencing data in 74/80 (93%) of these uveal melanomas (**Figure 2C and Supplementary Table 3**).

Of note, depending on the number of informative SNPs and their clonality in individual tumours, regional allelic imbalances involving other chromosomes were also successfully identified by this approach (**Figure 2A and B**).

### RNA-based identification of Gα_q_ and BSE mutations

Next, we aimed to identify the mutational status of individual uveal melanomas at the transcriptional level (**Figure 3A and Supplementary Table 3**). Gα_q_ signalling mutations are known to occur at a selected number of hotspots (p.G48, p.R183 and p.Q209 for *GNAQ* and *GNA11*, p.L129 for *CYSLTR2* and p.D630 for *PLCB4* [4-7]), allowing for a targeted analysis of RNA sequencing data (**Figure 3B**). Though, due to a lack of sequencing reads covering the mutant positions, DNA-confirmed *GNAQ* mutations could not be adequately detected in the RNA data of four cases (**Supplementary Figure 2A**). Still, the Gα_q_ mutations were correctly identified in the remaining 74/78 (95%) mutant uveal melanomas (**Figure 3A and Supplementary Table 3**).

**Figure 3.**
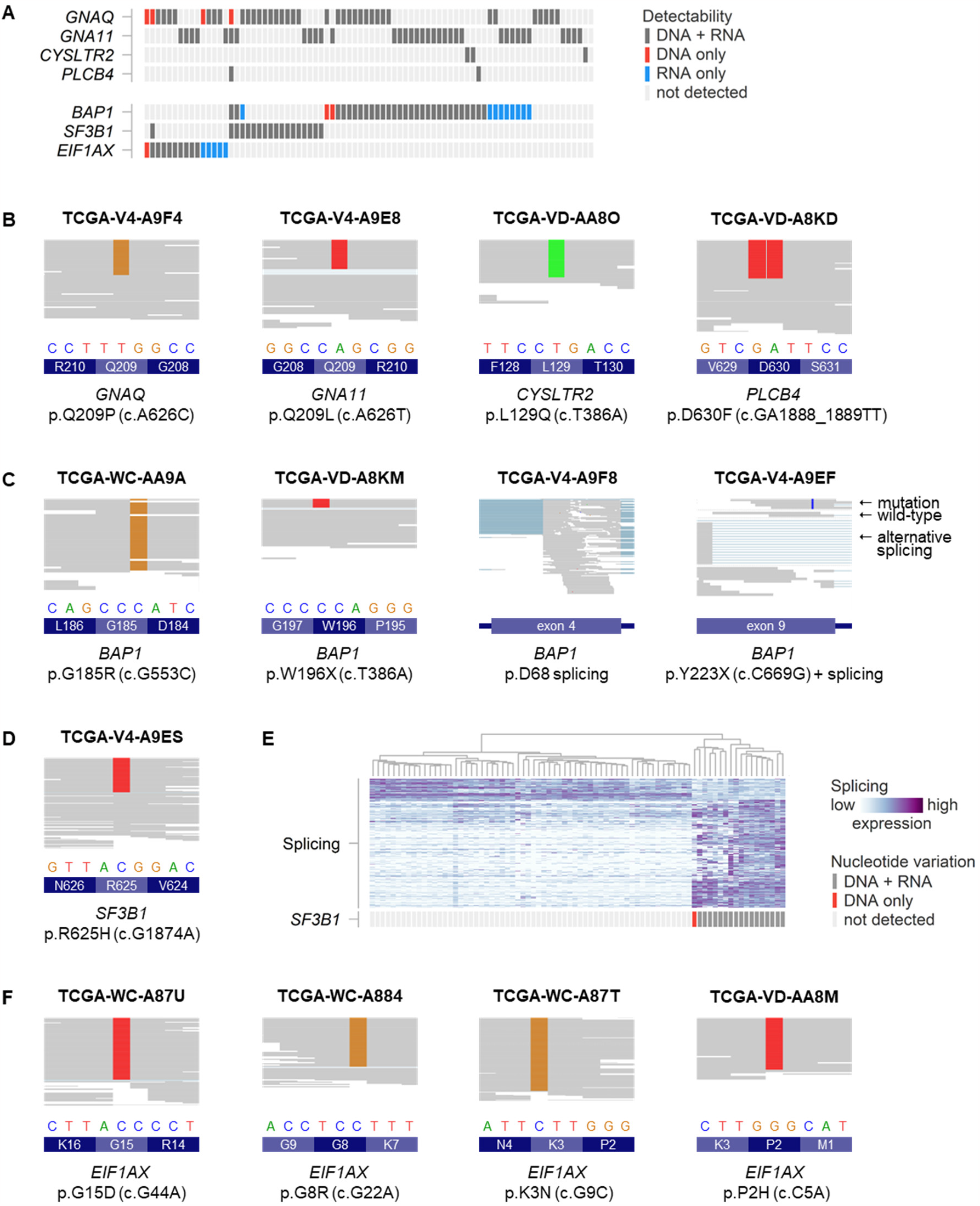
Detectability of mutations via RNA sequencing data in our training cohort of uveal melanomas studied by TCGA (n=80). (**A**) Summary of detected mutations in comparison to findings in exome-captured DNA sequencing. (**B**) Examples of detected Gα_q_ signalling mutations in RNA sequencing data. (**C**) Examples of detected *BAP1* mutations in RNA sequencing data: although some were identified directly via nucleotide variation, others showed alternative splicing around the mutant position. (**D**) Example of detected *SF3B1* mutation in RNA sequencing data. (**E**) Analysis of a signature of alternative splicing to identify *SF3B1* mutant tumours. (**F**) Examples of detected *EIF1AX* mutations in RNA sequencing data.

*BAP1* alterations are typically inactivating mutations (or deletions) that can be present throughout the complete *BAP1* gene. This challenges the identification of all alterations using conventional sequencing [25, 26]. In the TCGA cohort of uveal melanomas, it was earlier shown that various complex (intronic) *BAP1* alterations can be identified by the use of RNA sequencing [21, 25]. Importantly, exonic missense and nonsense mutations are also detectable at the transcriptional level: directly via their nucleotide changes, or indirectly via observed alternative splicing events such as (partial) intron retention or exon skipping (**Figure 3C**). Due to a low number of mutant RNA sequencing reads, two mutations were only identified at the DNA level (**Supplementary Figure 2A**). Taken together, 38/40 (95%) *BAP1* mutations could be detected via RNA sequencing (**Figure 3A and Supplementary Table 3**), including three mutations that had remained unrecognised in previous studies (p.W196X in VD-A8KM, a deletion involving the start of exon 1 in V4-A9ES, and a frameshift deletion involving exon 8 in WC-A883, **Figure 3C and Supplementary Figure 2B**).

*SF3B1* mutations in uveal melanomas are predominantly found in exon 14, with most of them affecting positions p.R625 or p.K666 [2, 9, 10, 24]. We successfully identified 14/15 (93%) DNA-confirmed mutations by nucleotide variation in the RNA sequencing data of the TCGA cohort (**Figure 3A, 3D and Supplementary Table 3**), with again a limited number of reads at the mutant position in the discordant tumour (**Supplementary Figure 2A**). However, alternative splicing was found in all 15/15 (100%) mutant tumour samples (**Figure 3E**), corroborating the differences between *SF3B1* mutant and wild-type uveal melanomas at the RNA level [24].

*EIF1AX* mutations mainly affect the N-terminal region of the gene, spanning the first two exons with a total of fifteen amino acids [10]. In the TCGA dataset, we detected the presence of a DNA-confirmed *EIF1AX* mutation in the RNA of 9/10 tumours, with – similar to the missed *GNAQ* mutation in the same tumour – a lack of sequencing reads explaining the discordant sample (**Supplementary Figure 2A**). However, we also observed that there was practically no coverage of exon 1 in the exome-captured DNA sequencing data, and none of the TCGA tumours was known to have an exon 1 mutation [21]. By analysing the (uncaptured) RNA sequencing data (**Figure 3D**), we identified expressed exon 1 mutations in five TCGA cases (p.P2H in VD-AA8M, p.P2L in YZ-A983, p.P2R in V4-A9EH, p.K3N in WC-A87T, and p.N4S in VD-AA8Q). All these tumours further presented with disomy of chromosomes 3 and 8q and lacked mutations in *SF3B1* or *BAP1*, in line with the typical genotype of *EIF1AX*-mutant tumours. For two cases, low-pass whole genome sequencing was available and confirmed both mutations (**Supplementary Figure 2C**). Taken together, 14/15 (93%) *EIF1AX* mutations were successfully identified at the RNA level, including five previously missed with DNA-based exome sequencing (**Figure 3A and Supplementary Table 3**).

### Prospective validation of RNA-based detectability of genetic alterations

Finally, we aimed to prospectively validate all beforementioned techniques in an independent cohort. For this goal, we performed RNA sequencing of five primary uveal melanoma specimens obtained after enucleation in our hospital (**Figure 4 and Supplementary Table 4**). The presence of chromosome 3 and 8q copy number alterations, and Gα_q_ and BSE mutations was confirmed at the DNA level using digital PCR and targeted sequencing, and – for BAP1 – at the protein level using immunohistochemistry.

**Figure 4.**
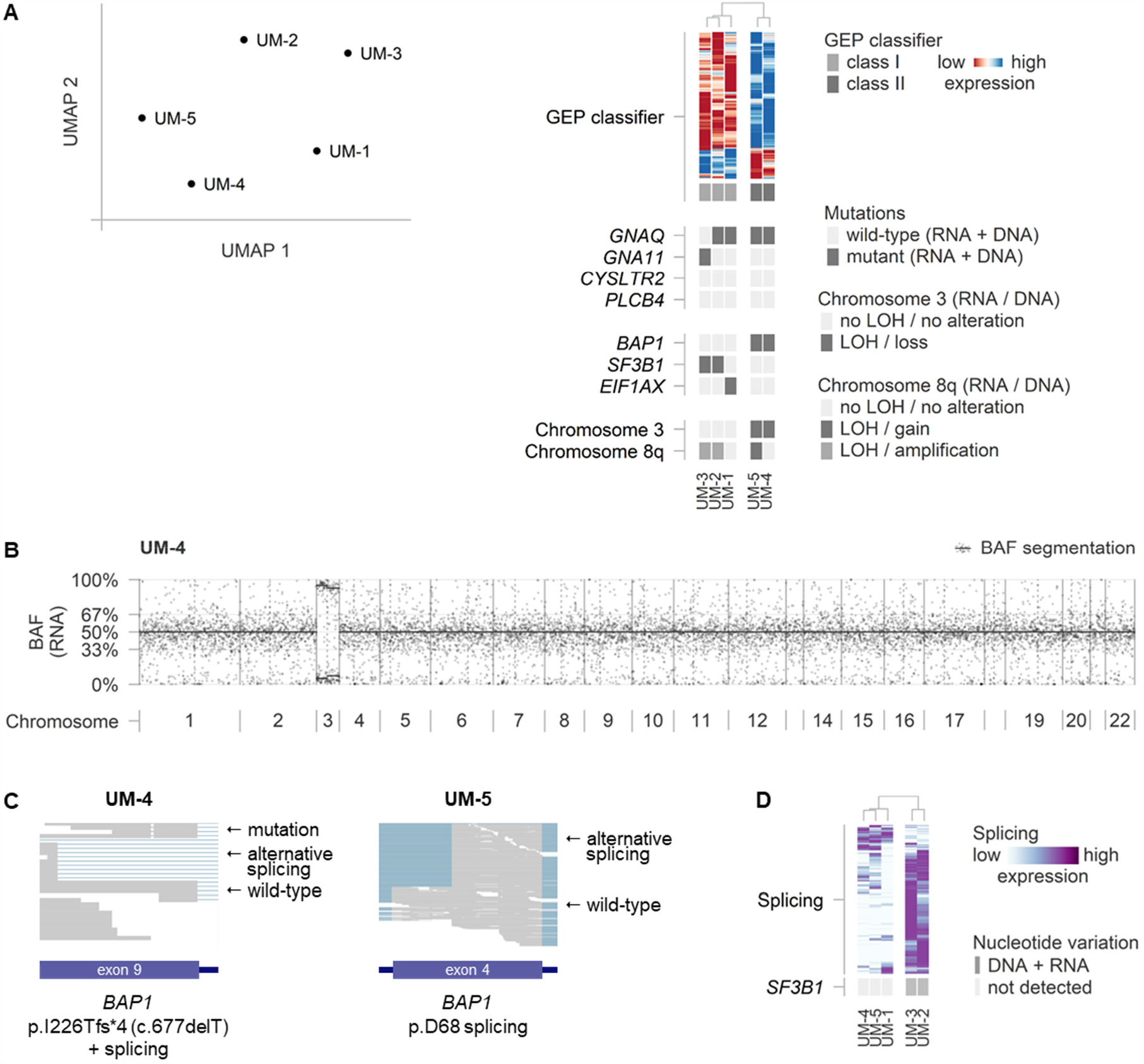
Analysis of our prospective cohort of uveal melanomas (n=5). (**A**) Two-dimensional uniform manifold approximation and projection (UMAP) analysis of cohort-wide gene expression profiling (GEP) and application of TCGA-derived GEP-classifier in relation to the RNA- and DNA-based detectability of genetic alterations. (**B**) Representative example of RNA-inferred allelic (im)balances to identify chromosomal alterations. In this tumour, imbalanced expression of genes of chromosome 3 correctly indicates the presence of a copy number alteration affecting this chromosome. In contrast, no imbalance is observed on chromosome 8q, in line with a disomy 8q. (**C**) *BAP1* alterations (mutations and mutation-associated alternative splicing) as observed in the RNA sequencing data. (**D**) Analysis of a signature of alternative splicing to identify *SF3B1* mutant tumours.

Although the number of tumours in our cohort was too low to reliably determine the GEP classification based on unsupervised clustering (**Figure 4A**), three tumours were found to be GEP class I and two tumours were GEP class II upon evaluation of our TCGA-based 200-gene expression signature. This classification matched with the loss of heterozygosity of chromosome 3 in the RNA sequencing data (exemplified in **Figure 4B**). Additionally, three tumours showed allelic imbalances of chromosome 8q. The presence of all these presumed copy number alterations could be successfully validated at the DNA level.

Regarding the Gα_q_ mutations, all five tumours expressed one hotspot mutation in *GNAQ* (n=4) or *GNA11* (n=1, **Figure 4B**). Next, the RNA data demonstrated the presence of BSE mutations in *BAP1* (n=2), *SF3B1* (n=2) and *EIF1AX* (n=1). The *BAP1* mutations caused RNA-detectable nucleotide variation as well as altered splicing around the mutated sites, showing high similarities to TCGA tumours with comparable mutations (**Figure 4C**). The tumours with the *SF3B1* p.R625 hotspot mutations were the ones demonstrating *SF3B1*-associated aberrant splicing (**Figure 4D**). One tumour carried an *EIF1AX* mutation affecting exon 2. All mutations could be verified at the DNA level. Additionally, the presence and pathogenicity of the *BAP1* alterations was supported by a negative (n=2) or positive (n=3) BAP1 nuclear staining (**Supplementary Table 4**).

Taken together, all measured copy number alterations and mutations were correctly identified in the RNA sequencing data of the five uveal melanomas.

## Discussion

Uveal melanoma is characterised by a number of prognostically-relevant combinations of genetic alterations, that are mostly measured at the DNA level. In this study, we tested whether these alterations could also be detected at the transcriptional level using RNA sequencing data. This identification of mutations and copy number alterations was cross-validated with DNA-based techniques in both a retrospective cohort of 80 TCGA tumours and five prospectively analysed uveal melanomas from our own cohort.

First of all, we explored the indirect detectability of genetic alterations via unsupervised GEP clustering, a commonly-used procedure in the context of classifying uveal melanoma. Our analysis revealed – in line with earlier studies – two main groups of tumours which overlapped with the absence or presence of chromosome 3 alterations (complete chromosomal loss and *BAP1* mutation, **Figure 1 and 4A**). Indeed, a large number of genes on chromosome 3 was expressed systematically lower in bulk RNA upon loss of this chromosome. However, GEP clustering was influenced by the extent of chromosome 3 loss (i.e. partial losses were not identified) and could not be used to specify the exact *BAP1* mutation. It also did not identify chromosome 8q copy number alterations or any of the other recurrent and relevant mutations, such as those in *SF3B1* and *EIF1AX*. As a further drawback, the GEP approach intrinsically relies on differences observed within a cohort, between multiple tumours. This makes this analysis less applicable to separate tumours, as it always requires a comparison to other samples (or ‘normal’ values) to identify any variation. For this reason, we further focussed on alternative approaches applicable to data of individual uveal melanomas.

To better identify chromosomal alterations, we measured regionally-abundant allelic imbalances of heterozygous common SNPs detectable in RNA data (**Figure 2**). This approach has several similarities to a DNA-based methodology we presented and discussed recently [31]. Based on (partial) copy number losses of chromosome 3, expressed loss of heterozygosity could be observed at the transcriptional level and turned out to be an excellent marker for this genetic event. The other way round, chromosome 8q copy number increases, predominantly mono-allelic gains or amplifications, were also readily detected by allelic imbalances present in RNA. Advantageously, by selecting SNPs known to be frequently heterozygous in the population, no patient-matched genotype information was required.

To detect BSE mutations at the RNA level, all mutational hotspot regions were screened and possible transcriptional consequences of various alterations (i.e. alternative splicing) were evaluated (**Figure 3**). Although a small number of mutations were missed due to insufficient sequencing coverage, the large majority was successfully detected in the RNA data. Interestingly, this included three additional *BAP1* mutations and five *EIF1AX* mutations in the TCGA cohort that were previously unrecognised as part of the respective genes were not or not sufficiently covered by the exome-captured DNA sequencing. Clinically, these alterations are prognostically relevant and not to be missed in any routine mutational screening of a uveal melanoma, substantiating complete coverage in any (targeted) sequencing panel. It is also essential that these alterations are known and being taken into account when analysing mutation data from these TCGA tumours in a research setting.

Though most mutations were directly recognised from nucleotide changes visible in the RNA sequencing reads, a number of *BAP1* mutations were identified via unusual splicing events within this gene (**Figure 3C and 4C**). In some tumours, the splice site disruption may be directly caused by the mutation being located near a regular exon-intron boundary, or the mutation introducing a novel splice site. Other tumours showed evidence of ‘nonsense-associated alternative splicing’: (partial) exon skipping or intron retention, apparently in response to the premature translation-termination codons introduced by nonsense or frameshift mutations. In contrast to nonsense-mediated decay, in-frame alternative splicing bypassing the original mutation might encode for a protein with saved functionalities, even though the mutation is predicted to be truncating and highly pathogenic [33, 34]. Our observations – possibly explainable by these phenomena, but yet to be followed up at a functional level – further contribute to the notorious complexity of *BAP1* alterations and their biological consequences [16, 17, 35-37].

Critically, our current study is limited by its low number of prospectively validated cases from independent cohorts. Still, the cross-validation in a total of 85 tumours (i.e. the TCGA and our own cohort combined) strongly suggests that most relevant alterations in uveal melanoma can be detected at the transcriptional level, especially when various bioinformatic approaches are combined. This indicates that RNA sequencing data, newly or retrospectively collected for gene expression analysis, may be mined for genetic alterations without the need to perform additional DNA-based measurements. Moreover, it may allow researchers to study genotype-phenotype relations in the same tumour specimen in more detail. This is particularly interesting in the context of measuring genetic heterogeneity next to transcriptional heterogeneity, for example using multiregional or single-cell RNA sequencing. Thirdly, the detectability of genetic alterations in the RNA of uveal melanomas may provide the rationale to study (cell-free) RNA from liquid biopsies, which already showed potential in other types of cancer [38-41].

In a follow-up study, our evaluations may be extended to other genetic alterations in uveal melanoma. Besides chromosome 3 and 8q, (partial) copy number alterations affecting chromosome 1, 6 and 16 are recurrently present and – although not extensively validated – were already correctly observed as allelic imbalances in some of our tumours (**Figure 2 and Supplementary Figure 1**). Additionally, mutations in splicing factor *SRSF2*, which seem to form a rare alternative to *SF3B1* mutations [21, 42], might be recognised in the RNA sequencing reads or by its own signature of alternative splicing. Moreover, it would be therapeutically relevant to evaluate alterations involving *MBD4*. Biallelic inactivation of this gene (via a mutation and copy number loss of its locus on chromosome 3) has been identified as a rare cause of CpG>TpG hypermutation in uveal melanoma, conferring unique sensitivity to immunotherapy [43, 44]. Next to identifying mutations at the RNA level, additional evidence for MBD4 inactivation can be found in an unusually high total mutational burden, which might also be inferred from RNA sequencing data [45]. Finally, our methodologies might be valuable beyond the field of uveal melanoma, and it would be interesting to evaluate these in other types of cancer.

In conclusion, we developed and applied a variety of bioinformatic approaches showing that transcriptional data – in addition to providing gene expression levels and profiles – forms a rich source of (clinically-)relevant genetic information in uveal melanoma. Via RNA, valuable insights could be obtained into the expressed genotype and its phenotypic consequences, but this also demonstrated the vulnerabilities of an (insufficiently) targeted DNA measurement. As illustrated by the identification of previously unreported mutations in the thoroughly studied TCGA cohort, and the successful prospective analysis of tumours of our own cohort, the analysis of transcriptional data may augment or even substitute current DNA-based approaches, and has potential applicability in both oncological research and clinical practice.

## Data Availability

This work is partly based upon data generated by The Cancer Genome Atlas (TCGA) Research Network: https://www.cancer.gov/tcga. All data produced in the study are contained in the manuscript and supplement.

https://github.com/rjnell/um-rna

## Acknowledgements

This study is supported by the European Union’s Horizon 2020 research and innovation program under grant agreement number 667787 (UM Cure 2020, Rogier J. Nell). We thank Ronald van Eijk (Department of Pathology, Leiden University Medical Center, Leiden, the Netherlands) for his help with the *BAP1* DNA sequencing experiments. This work is partly based upon data generated by The Cancer Genome Atlas (TCGA) Research Network: https://www.cancer.gov/tcga.

