## Supplementary Information for "Identification of clinically-relevant genetic alterations in uveal melanoma using RNA sequencing"

### Supplementary Table 1

Context sequence and annealing temperature for novel digital PCR assays used in this study.

| Assay | *SF3B1* p.R625 mutation assay |
|---|---|
| Context sequence | GGTCTGGCTA CTATGATCTC TACCATGAGA CCTGATATAG ATAACATGGA TGAGTATGTC CGTAACACAA CAGCTAGAGC TTTTGCTGTT GTAGCCTCTG CCCTGGGCAT TCCTTCTTTA TTG |
| Location (hg38) | chr2:197402698-197402820 |
| Amplicon length | 77 nucleotides |
| Annealing temperature | 55 °C |

| Assay | *EIF1AX* exon 2 mutation assay |
|---|---|
| Context sequence | TTATGAAAAC ACTTACCCTG ACCATCCTCT TTGAATACCA GTTCTCTTTT TTCAGATTCA TTCTCATTCT TACCCCTGCG TCTGTTTTTA CCTCCTTTAC CTGATGGTTT AAAAAAAAGA AAAGGAGGTA AATGACATTA ATTATCTGTA AAACAGTATG AAATAAAATG AAATTAA |
| Location (hg38) | chrX:20138523-20138699 |
| Amplicon length | 123 nucleotides |
| Annealing temperature | 58 °C |

### Supplementary Table 2

Details of the inferred signature of 200 genes characterising the two clusters identified by unsupervised clustering of transcriptome-wide expression data.

### Supplementary Table 3

Overview of genetic and transcriptional findings in our retrospective cohort of 80 uveal melanomas studied by The Cancer Genome Atlas (TCGA).

### Supplementary Table 4

Overview of genetic, transcriptional and immunohistochemical findings in our prospective cohort of five uveal melanomas.

**Supplementary Figure 1**

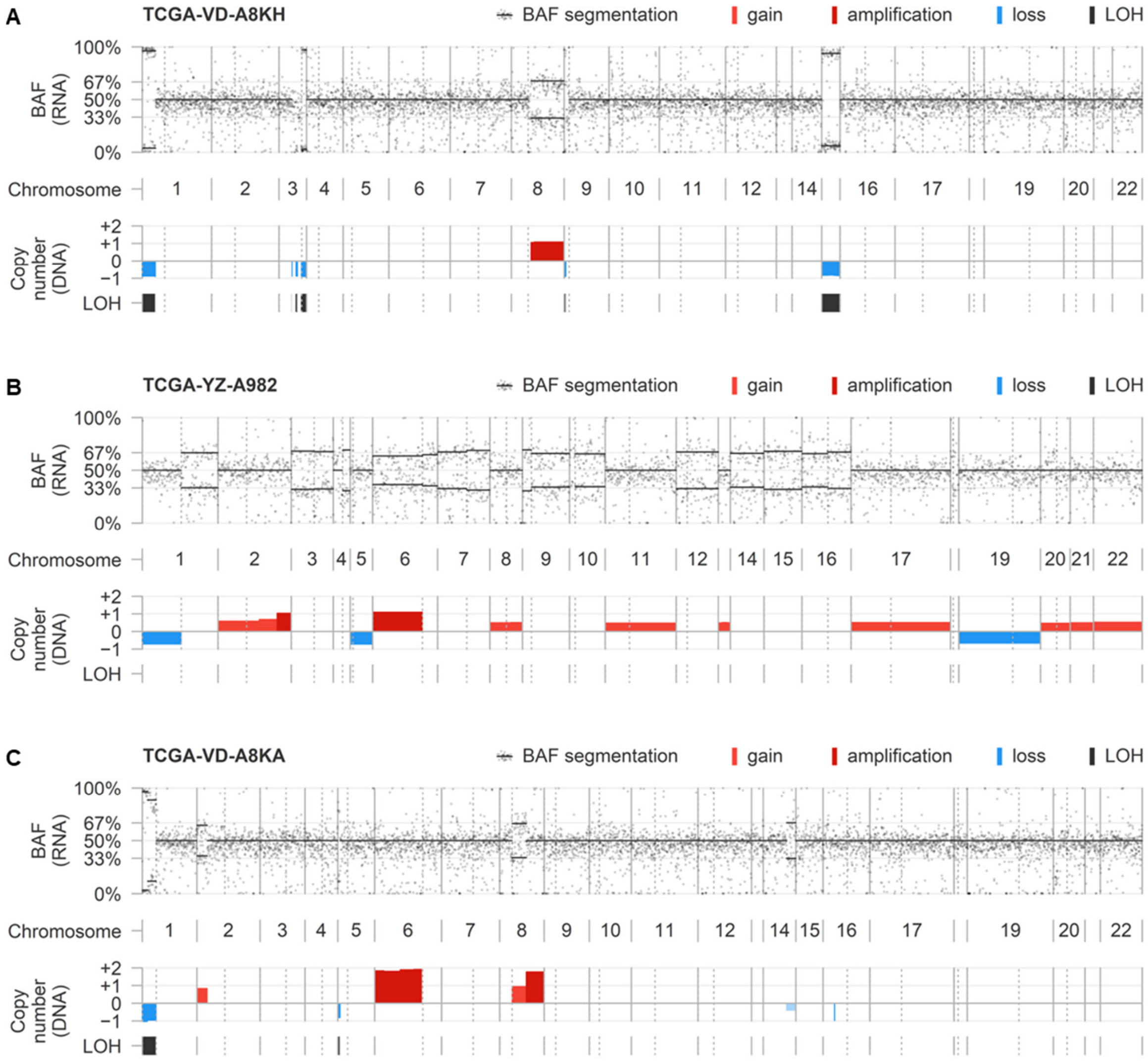


RNA-inferred allelic imbalances in relation to DNA-confirmed copy number alterations.

(**A**) Example of a tumour showing variation in the expressed BAFs on chromosome 3: chromosome 3p is largely balanced in line with a disomy, but 3q demonstrates imbalances in the context of a monosomy. Additionally, alterations affecting chromosome 1p, 8q, 9p and 15 are correctly observed.

(**B**) Example of a tumour showing imbalanced RNA expression and DNA-measured copy number alterations affecting the majority of chromosomes. A polyploid genotype (most likely n=3) is suggested as the copy number neutral state is associated with BAFs close to 33% and 67%, and relative losses/gains with BAFs close to 50%.

(**C**) Example of a tumours showing DNA-confirmed amplifications affecting chromosomes 6p and 8q not associated with BAF deviations, likely because both amplifications are derived from biallelic genetic events.

**Supplementary Figure 2**

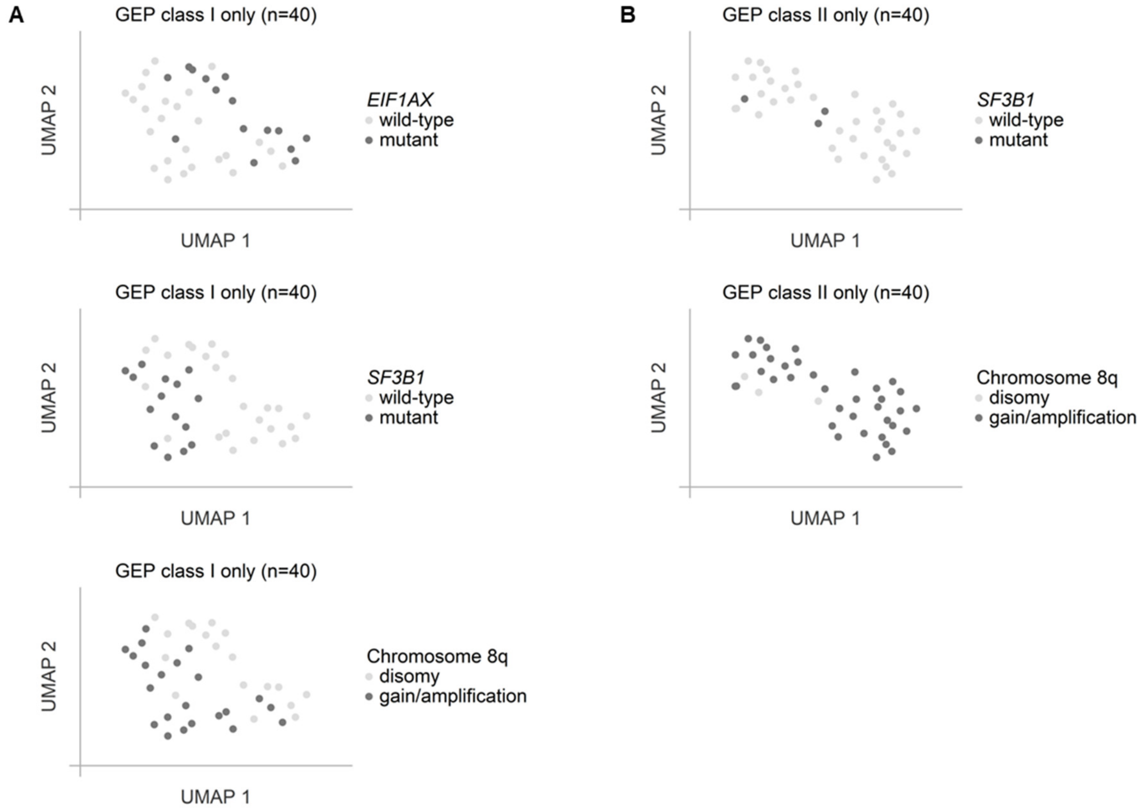


Two-dimensional uniform manifold approximation and projection (UMAP) analysis, specified to TCGA tumours within the class I and II gene expression profiling (GEP) clusters.

(**A**) Distribution of *EIF1AX* mutations, *SF3B1* mutations and chromosome 8q copy number alterations across GEP class I tumours.

(**B**) Distribution of *SF3B1* mutations and chromosome 8q copy number alterations across GEP class II tumours.

**Supplementary Figure 3**

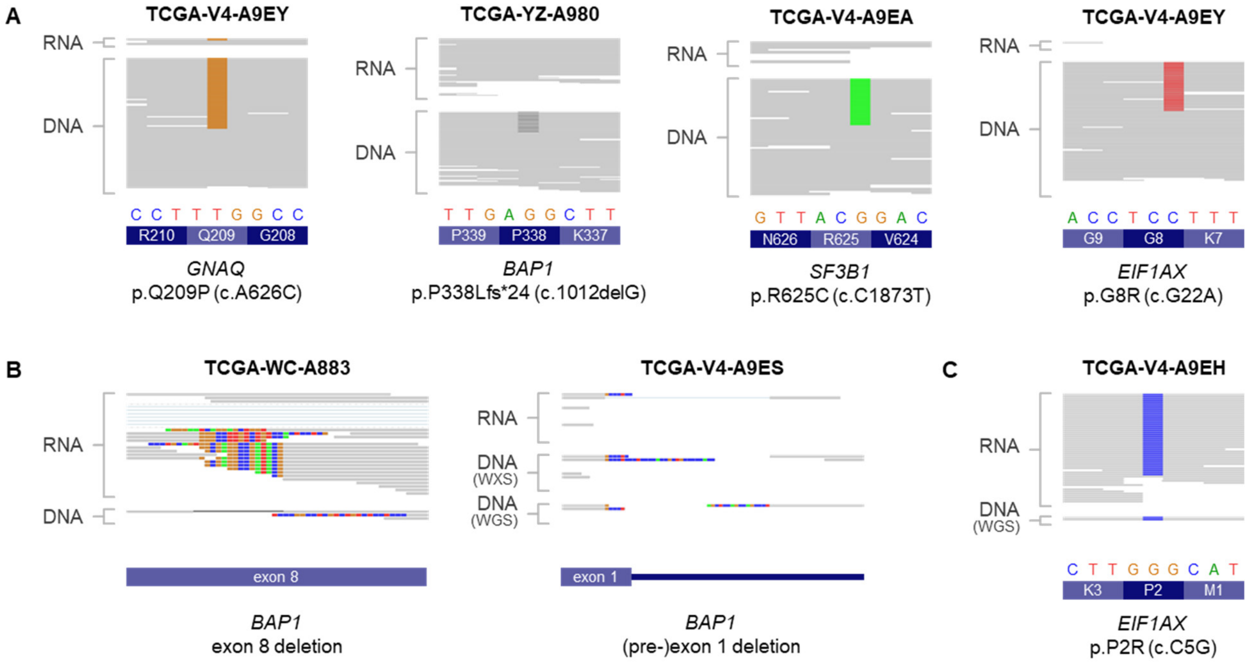


Detectability of mutations via RNA sequencing, in comparison to standard exome-captured DNA sequencing (WXS) or low-pass whole-genome sequencing (WGS, not available in all cases).

(**A**) Examples of mutations not detected in RNA data, due to a low number of total or mutant reads.

(**B**) Examples of *BAP1* mutations newly identified by analysing RNA data in addition to DNA sequencing. Note that automatic alignment failed in recognising the deletions.

(**C**) Example of a previously unrecognised *EIF1AX* exon 1 mutation, detected via RNA sequencing and confirmed by low-pass WGS.
